# Bucillamine use for rheumatoid arthritis and type 2 diabetes mellitus are associated with neural epidermal growth factor-like 1 (NELL1)-associated membranous nephropathy

**DOI:** 10.1101/2022.12.02.22282616

**Authors:** Takahiro Tsuji, Sari Iwasaki, Keishi Makita, Shota Furukawa, Kanako Watanabe-Kusunoki, Sayo Takeda-Otera, Takahito Itoh, Mamiko Shimamoto, Hiroaki Yamaji, Tomomasa Yoshimura, Junya Yamamoto, Takashi Kudo, Makoto Kondo, Hiroshi Kataoka, Masaya Mukai, Yukito Kaga, Miku Yoshinari, Yuka Nishibata, Sakiko Masuda, Utano Tomaru, Akihiro Ishizu, Yuichiro Fukasawa, Seiji Hashimoto, Saori Nishio

**Affiliations:** Department of Pathology, Sapporo City General Hospital, Hokkaido, Japan; Department of Nephrology, Sapporo City General Hospital, Hokkaido, Japan; Department of Rheumatology and Clinical Immunology, Sapporo City General Hospital, Hokkaido, Japan; Department of Clinical Laboratory, Sapporo City General Hospital, Hokkaido, Japan; Department of Pathology, Faculty of Medicine and Graduate School of Medicine, Hokkaido University, Hokkaido, Japan; Department of Surgical Pathology, Hokkaido University Hospital, Hokkaido, Japan; Department of Rheumatology, Endocrinology and Nephrology, Faculty of Medicine and Graduate School of Medicine, Hokkaido University, Hokkaido, Japan; Department of Medical Laboratory Science, Faculty of Health Sciences, Hokkaido University, Hokkaido, Japan; Department of Internal Medicine, Kushiro Red Cross Hospital, Hokkaido, Japan; Department of Cardiology, Oji General Hospital, Tomakomai, Hokkaido, Japan; Department of Pathology, Oji General Hospital, Tomakomai, Hokkaido, Japan; Department of Nephrology, Otaru General Hospital, Hokkaido, Japan; Tomakomai Urology and Cardiology Clinic, Hokkaido, Japan; Department of Nephrology, JCHO Hokkaido Hospital, Hokkaido, Japan; Department of Nephrology, Kinan Hospital, Tanabe, Wakayama, Japan

**Keywords:** Membranous nephropathy, NELL1, bucillamine, rheumatoid arthritis, type 2 diabetes mellitus

## Abstract

Membranous nephropathy (MN) is a disease characterized by deposition of immune complexes on the glomerular basement membrane. More than 10 specific antigens for MN including M-type phospholipase A2 receptor (PLA2R), thrombospondin type-1 domain-containing 7A (THSD7A), exostosin 1/exostosin 2 (EXT1/EXT2) and neural epidermal growth factor-like 1 (NELL1) have so far been identified. Since the clinicopathologic characteristics of each type of MN in Japanese are not well understood, we first examined 107 cases of MN by immunohistochemistry for four antigens (PLA2R, THSD7A, EXT1, and NELL1) (MN-cohort). Of those 107 cases, 40% were PLA2R-positive, 13% were NELL1-positive, 11% were THSD7A-positive, 5% were EXT1-positive, 2% were PLA2R and NELL1-double-positive, and 29% were quadruple-negative. In one case of PLA2R and NELL1-double-positive, the first biopsy showed PLA2R-positive and the second biopsy showed PLA2R and NELL1-double-positive. Of the 16 cases of NELL1-positive, 12.5% had colon cancer, 18.8% had rheumatic diseases treated with bucillamine, and 63% had type 2 diabetes mellitus (T2DM). Next, 34 patients diagnosed with MN who had rheumatoid arthritis (RA) were examined (RA-MN cohort). Of those 34 patients, 79% were NELL1-positive, 6% were PLA2R-positive, and 15% were quadruple-negative. In the RA-MN cohort, 56% had a history of bucillamine use. In conclusion, NELL1-associated MN is a common MN in patients with RA using bucillamine and may also be associated with T2DM.

## Introduction

Membranous nephropathy (MN) is defined histopathologically as a disease characterized by podocyte injury due to deposition of immune complexes, most of which contain immunoglobulin G (IgG), between the glomerular basement membrane and podocytes. Clinically, the disease is characterized by slowly progressive proteinuria and often presents with nephrotic syndrome. Historically, MN has been classified as primary (or idiopathic) MN, in which lesions are confined to the kidney, and secondary MN, which is a form of MN secondary to other causes such as malignancy, drugs, autoimmune diseases, or infections. Reports of specific antigens for MN began with the identification of M-type phospholipase A2 receptor (PLA2R) and thrombospondin type-1 domain-containing 7A (THSD7A).^1,2^ From 2019 onwards, the success of combined laser microdissection and mass spectrometry techniques led to the discovery of various antigens including exostosin 1/exostosin 2 (EXT1/EXT2) ^3^, neural epidermal growth factor-like 1 protein (NELL1) ^4^, semaphorin 3B ^5^, neural cell adhesion molecule 1 ^6^, protocadherin 7 ^7^, serine protease HTRA1 ^8^, transforming growth factor beta receptor 3 ^9^, protocadherin FAT1 ^10^, contactin 1 ^11^ and netrin G1 ^12^. As of 2022, it is estimated that the complete picture of the specific antigens of MN is close to being elucidated. In this study, we immunohistochemically examined four antigens, PLA2R, THSD7A, NELL1 and EXT1, in cases of MN for which biopsy specimens were collected mainly in Hokkaido, the northernmost island of Japan, and we determined their respective frequencies. We then focused on NELL1-associated MN, the second most common type of MN after PLA2R-associated MN, and conducted a clinicopathological study. In our study, NELL1-associated MN was frequently observed in patients with type 2 diabetes mellitus (T2DM) and rheumatoid arthritis (RA) who were using bucillamine (BUC) as a therapeutic agent in addition to patients with malignancies.

## Methods

### Patients and data collection

Patients with histopathologically diagnosed MN at Sapporo City General Hospital were included in this study. A diagnosis of MN is made when both of the following histologic criteria are met: 1) immunofluorescence shows granular deposition of IgG along the glomerular basement membrane and 2) light microscopy shows a membranous pattern of tissue injury such as pinhole or spike formation on the glomerular basement membrane by silver staining.

The Department of Pathology at Sapporo City General Hospital receives renal biopsy specimens from all over Hokkaido, the northernmost island of Japan. This study consisted of two cohorts (MN cohort and RA-MN cohort; Figure 1) and the study was approved by the Ethics Committee of Sapporo City General Hospital (Approval No. R02-060-759) MN cohort: Of 1169 renal biopsy cases diagnosed between May 2017 and September 2020, 107 cases of MN excluding lupus nephritis type V were included.

**Figure 1.**
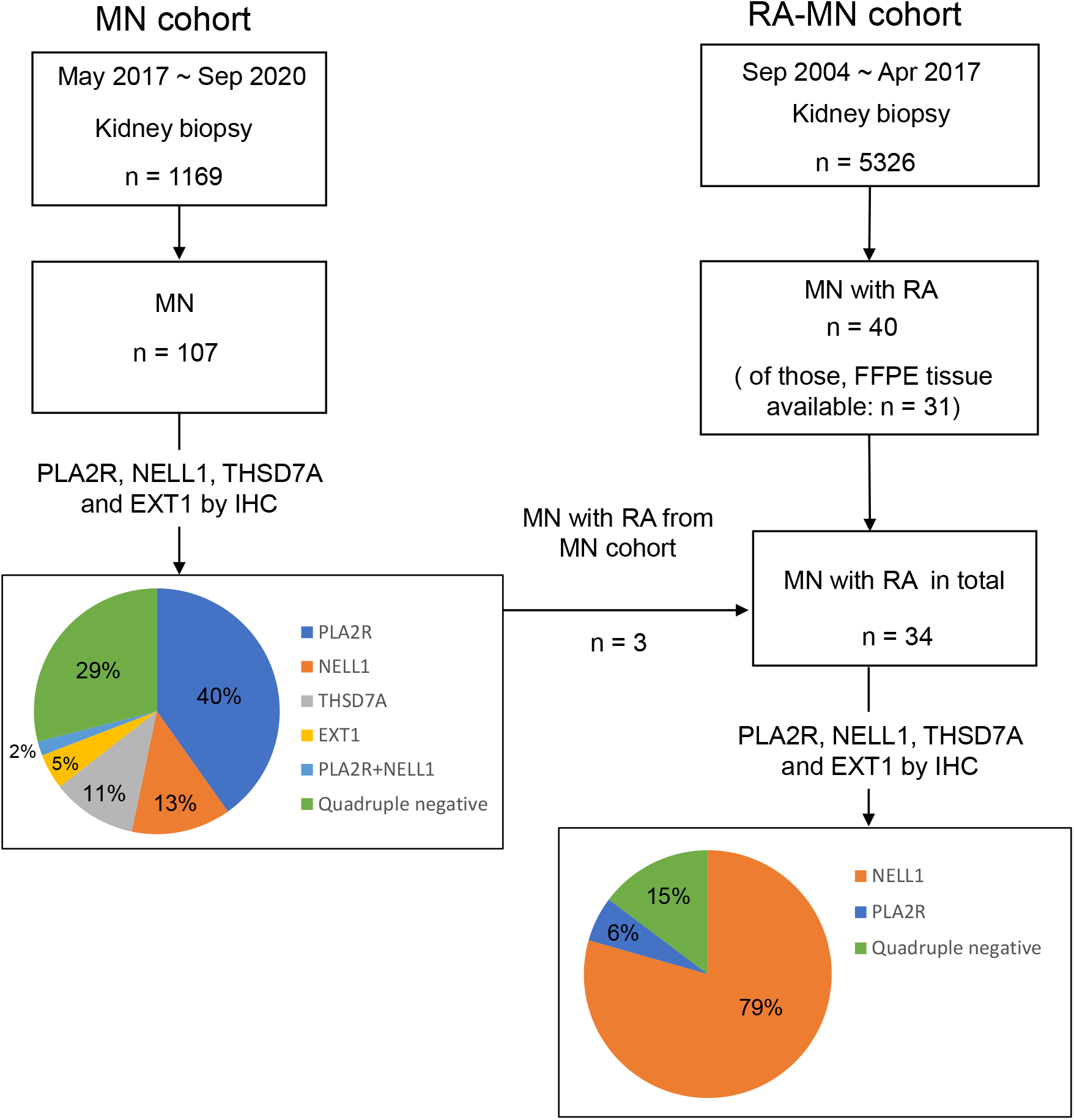
Study design of the MN cohort and the RA-MN cohort. In the MN cohort, 107 patients with membranous nephropathy (MN), excluding lupus nephritis type V, were studied by immunohistochemistry (IHC) for 4 antigens: M-type phospholipase A2 receptor (PLA2R), thrombospondin type-1 domain-containing 7A (THSD7A), neural epidermal growth factor-like 1 (NELL1), and exostosin 1 (EXT1). In the MN cohort, PLA2R was the most common antigen, accounting for 40% of the cases, and NELL1 was the second most common antigen, accounting for 13% of the cases. In addition, 3 cases of NELL1-associated MN in the MN cohort had rheumatoid arthritis (RA), which led us to plan a subsequent RA-MN cohort. In the RA-MN cohort, IHC was performed for 4 antigens, PLA2R, THSD7A, NELL1 and EXT1, in 34 patients including 31 patients with MN who had RA and 3 MN patients with MN with RA who were included in the MN cohort.

Clinical information at the time of biopsies including information on age, gender, serum creatinine level, serum albumin level, proteinuria, and presence of concomitant diseases such as malignancy and autoimmune diseases were obtained from the kidney biopsy order form. For pathological information, findings of light microscopy, immunofluorescence, immunohistochemistry (IHC) for 4 antigens (PLA2R, THSD7A, NELL1 and EXT1), and electron microscopy were collected. For cases of NELL1-associated MN, information on treatment and follow-up was also obtained from clinicians.

RA-MN cohort: Among 5326 renal biopsy cases diagnosed between September 2004 and September 2017, 31 cases of MN with RA for which formalin-fixed paraffin-embedded tissue (FFPE) was available were included. The RA-MN cohort also included three cases of NELL1-associated MN with RA from the MN cohort. Clinical information at the time of biopsies including information on age, gender, serum creatinine level, serum albumin level, and proteinuria as well as types of anti-rheumatic drugs and presence of comorbidities was obtained from the request forms for renal biopsies. IHC for PLA2R, THSD7A, NELL1, and EXT1 was performed using FFPE tissue.

## Results

### NELL1 was the second most common antigen in the MN cohort

In the MN cohort, IHC for four MN-specific antigens (PLA2R, THSD7A, NELL1, and EXT1) was performed in specimens from all patients (n=107) diagnosed with MN at our institution between May 2017 and September 2020 (Figure 1).

Forty-three cases (40%) were positive for PLA2R, 14 cases (13%) were positive for NELL1, 12 cases (11%) were positive for THSD7A, 5 cases (5%) were positive for EXT1, 2 cases (2%) were double-positive for PLA2R and NELL1, and 31 cases (29%) were quadruple-negative (Figure. 1). Therefore, we decided to focus on NELL1-associated MN, which was the second most frequent, and proceeded with subsequent clinicopathological studies.

### Clinicopathological characteristics of NELL1-associated MN (MN cohort)

The clinical characteristics and pathological characteristics of NELL1-associated MN found in the MN cohort are shown in Tables 1 and 2, respectively. A total of 16 cases of NELL1-associated MN were identified. The 16 cases included 14 cases that were NELL1-positive alone (cases 1 through 14) and 2 cases that were PLA2R- and NELL1-double positive (cases 15 and 16). Clinical information at the time of renal biopsy is shown in Table 1. The male-to-female ratio of patients with NELL1-associated MN was 11:5 and the mean age of the patients was 74.6 years (standard deviation (SD)±7.8). The mean serum creatinine level was 1.1 mg/dl (SD±0.6), the mean serum albumin level was 2.4 g/dl (SD±0.9), and the mean urinary protein level was 7.0 g/gCr (SD±5.2) or 7.1 g/day (SD±4.7). Two patients (12.5%: case 3 and case 13) had malignancies, both of which were colon cancer. Five patients had rheumatic diseases (31.3%: RA in 3 cases (cases 11-13), polymyalgia rheumatica (PMR) in 1 case (case 10), and spondyloarthritis (SpA) in 1 case (case 14)). Other complications were asthma (case 1), cellulitis (case 2), benign prostatic hyperplasia (case 15), and recurrent MN (case 16). T2DM was noted in 10 patients (63%: cases 2-4, 6, 7, 9, 12, 13, 15, 16). In 8 cases of T2DM in which we were able to confirm a history of insulin therapy, there was no history of insulin therapy.

**Table 1.** Clinical characteristics of NELL1-associated membranous nephropathy found in the MN cohort

| Clinical information at the time of biopsy |  |  |  |  |  |  |  |  |  | Treatment and clinical follow up |  |  |  |
| --- | --- | --- | --- | --- | --- | --- | --- | --- | --- | --- | --- | --- | --- |
| Case | Age<sup>a</sup><br>( years ) | Sex | Serum<br>Cr,<br>mg/dl | Serum<br>albumin,<br>g/dl | Urinary<br>protein,<br>g/gCr | Urinary<br>protein,<br>g/24 h | HbA1c, % | Insulin<br>therapy | Other diseases | Treatment | Observation<br>period, months | Serum Cr<br>at last<br>follow-up,<br>mg/dl | Urinary<br>protein at<br>last follow-up, g/gCr |
| 1 | 86-90 | M | 1.6 | 2.0 | n.a. | 5.0 | n.a. |  | asthma | PSL | n.a. | n.a. | n.a. |
| 2 | 71-75 | F | 0.6 | 1.4 | 17.7 | n.a. | 6.2 | - | T2DM, cellulitis | ARB | 31 | 0.67 | 0.32 |
| 3 | 76-80 | M | 3.0 | 2.1 | 0.8 | 0.0 | 5.9 | n.a. | Colon cancer,<br>T2DM | n.a. | n.a. | n.a. | n.a. |
| 4 | 71-75 | M | 0.8 | 2.8 | 6.7 | 2.7 | 6.9 | - | T2DM | PSL, CyA,<br>mPSL | 26 | 1.19 | 0.12 |
| 5 | 66-70 | F | 0.7 | 3.5 | 5.8 | n.a. | 5.5 |  | - | ARB | 25 | 0.8 | 0.04 |
| 6 | 76-80 | F | 1.3 | 3.2 | 5.3 | n.a. | 7.7 | - | T2DM | no treatment | 6 | 1.08 | 2.4 |
| 7 | 81-85 | M | 1.0 | 2.6 | n.a. | 11.4 | 6.3 | - | T2DM | PSL, CyA | 4 | 0.91 | 1.66 |
| 8 | 86-90 | M | 1.0 | 1.8 | n.a. | 8.0 | n.a. |  | - | PSL, CyA | 8 | 1.05 | 4.76 |
| 9 | 66-70 | M | 0.9 | 1.1 | n.a. | 12.8 | 7.1 | - | T2DM | PSL, CyA,<br>ARB | 3 | 0.7 | 0.17 |
| 10 | 76-80 | F | 0.6 | 3.3 | 7.7 | n.a. | 5.7 |  | PMR | PSL, ARB | 4 | 0.76 | 11.16 |
| 11 | 66-70 | M | 0.8 | 0.7 | 10.2 | 9.7 | n.a. |  | RA | BUC -> IGU | 7 | 0.77 | 1.84 |
| 12 | 71-75 | M | 1.3 | 3.3 | n.a. | 4.4 | n.a. | - | RA, T2DM | BUC -> AZA | 4 | 1.13 | 2.8 |
| 13 | 56-60 | F | 1.1 | 1.7 | n.a. | 12.7 | 6.1 | - | Colon cancer, RA,<br>T2DM | Colon EMR,<br>MTX -> IGU | 1 | 1.04 | 1.74 |
| 14 | 66-70 | M | 0.8 | 4.1 | 0.9 | 0.5 | n.a. |  | SpA | BUC -> mPSL,<br>TAC | 1 | 0.81 | 0.18 |
| 15 | 76-80 | M | 0.8 | 2.4 | 12.1 | n.a. | 6.9 | n.a. | T2DM, prostatic<br>hyperplasia | PSL, CyA | 11 | 1.23 | 1.33 |
| 16 | 66-70 | M | 1.1 | 2.2 | 3.4 | 6.0 | 6.7 | - | T2DM, recurrent<br>MN | ARB | 1 | 1.26 | 3.94 |

Abbreviations: ARB, angiotensin II receptor blocker; AZA, azathioprine; BUC, bucillamine; Cr, creatinine; CyA, cyclosporine A; EMR, endoscopic mucosal resection; F, female; HbA1c, hemoglobin A1c; IGU, iguratimod; M, male; MN, membranous nephropathy; mPSL, methylprednisolone; MTX, methotrexate; n.a., not available; NELL1, neural epidermal growth factor-like 1; PMR, polymyalgia rheumatica; PSL, prednisolone; RA, rheumatoid arthritis; SpA, spondyloarthritis; T2DM; type 2 diabetes mellitus
<sup>a</sup>Patient ages are listed with a range of 5 years to protect personal information.

**Table 2.**
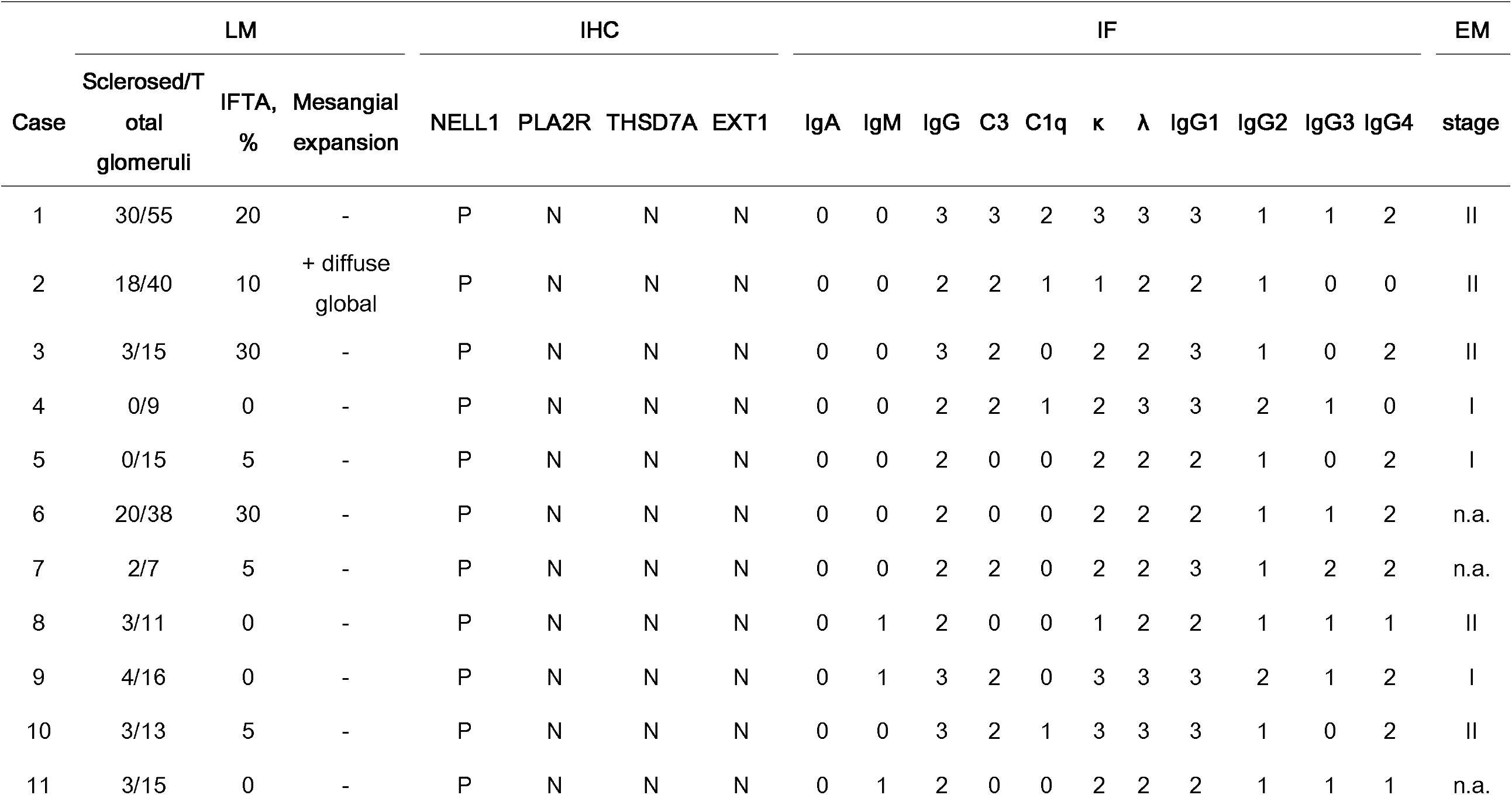
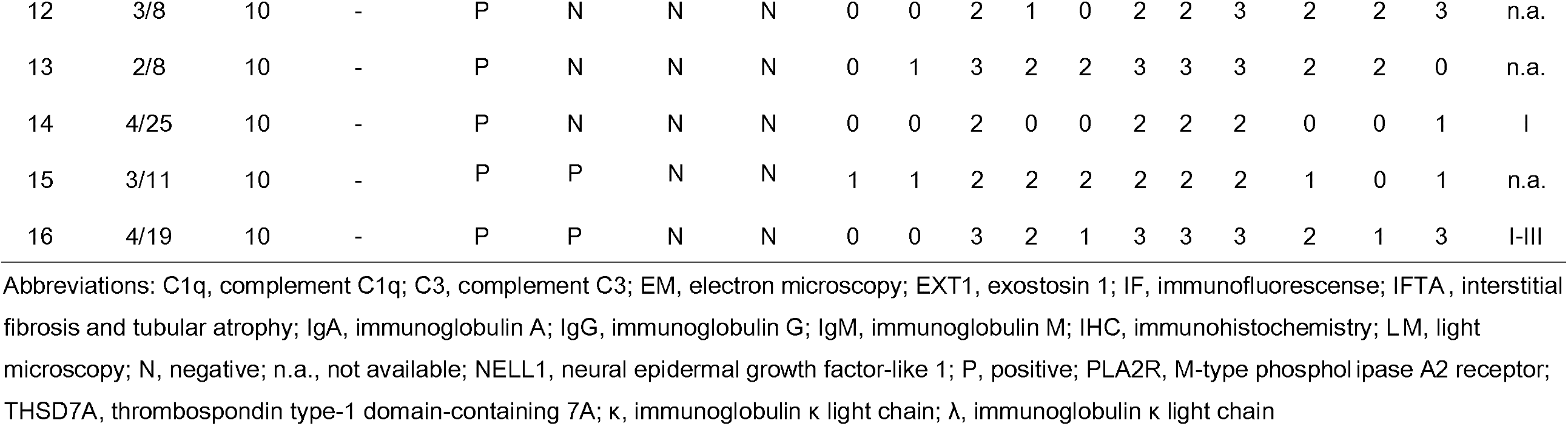
Pathological features of NELL1-associated membranous nephropathy found in the MN cohort

Treatment information and follow-up information are shown on the right side of Table 1. Five patients received only immunosuppressive therapy with prednisolone and/or cyclosporine A (CyA) (cases 1, 4, 7, 8, 15), two patients received both immunosuppressive therapy and angiotensin receptor blockers (cases 9, 10), and three patients received angiotensin receptor blockers only (cases 2, 5, 16). One patient (case 6) was followed up with no treatment. In four patients with RA or spondyloarthritis, drug-induced MN due to anti-rheumatic drugs was suspected, and the drug was changed (bucillamine (BUC): cases 11, 12, 14; methotrexate (MTX): case 13). Endoscopic mucosal resection of colon cancer was performed in one case (case 13). The mean observation period was 9.4 months (SD ± 10.2), the latest serum creatinine level was 1.0 mg/dl, and the latest urinary protein level was 2.3 g/gCr.

The pathological characteristics of the renal biopsy specimens are shown in Table 2. On average, 19.1 (SD ± 13.8) glomeruli were included, of which 6.4 (SD ± 8.5) were globally sclerotic. The IFTA area averaged 9.7% (SD ± 9.6). Diffuse global mesangial expansion with mesangial cell proliferation was observed in one case with T2DM (case 2), which was considered to be complicated with diabetic nephropathy Class IIa ^13^. The remaining 9 patients who had T2DM did not show mesangial expansion and were not considered to have diabetic nephropathy.

Immunofluorescence showed clear IgG positivity of moderate or strong (2-3+) in all cases. IgA and IgM were not significantly positive in any case. C3 was moderately or strongly positive (2-3+) in 10 patients (63%). C1q was moderately (2+) positive in 3 cases (19%). There was no significant deviation in the deposition of immunoglobulin light chains. Analysis of IgG subclasses showed that all patients were IgG1 dominant (100%). Of these, IgG4 was co-dominant in 5 cases (31%). IgG2 and IgG3 were moderately positive (2+) in 5 cases (31%) and 3 cases (19%), respectively, but the intensity of positivity was weaker than that of IgG1 in all cases.

Electron microscopic analysis was performed in 10 cases, and 4 were classified as stage I, 5 were classified as stage II, and 1 was classified as stage I-III according to Ehrenreich and Churg’s classification.

### Analysis of MN with RA (RA-MN cohort)

The results of the MN cohort showed that NELL1-associated MN was suspected to be associated with RA and anti-rheumatic drug use, in addition to malignancy and T2DM. Therefore, we planned the RA-MN cohort using 34 cases, including 31 cases with previously diagnosed MN with RA and 3 cases of MN with RA in the MN cohort (Figure 1, Table 3).

**Table 3.** Clinical characteristics and immunohistochemistry results in the RA-MN cohort cases

| Case | Clinical information at the time of biopsy |  |  |  |  |  |  | Immunohistochemistry |  |  |  |
| --- | --- | --- | --- | --- | --- | --- | --- | --- | --- | --- | --- |
|  | Age<sup>b</sup><br>(years) | Sex | Serum<br>Cr, mg/dl | Serum<br>albumin,<br>g/dl | Urinary<br>protein,<br>g/gCr | anti-rheumatic drugs | Other diseases | NELL1 | PLA2R | THSD7A | EXT1 |
| 1 | 66-70 | F | 0.6 | 2.0 | 14.1 | BUC (WD) | HCV(+) | P | N | N | N |
| 2 | 71-75 | F | 1.1 | 2.5 | 3.6 | BUC (WD) |  | P | N | N | N |
| 3 | 36-40 | F | 0.7 | 3.2 | n.a. | BUC (WD) |  | P | N | N | N |
| 4 | 61-65 | F | 0.6 | 2.1 | 4.5 | BUC |  | P | N | N | N |
| 5 | 61-65 | F | 0.6 | 1.8 | n.a. | BUC |  | P | N | N | N |
| 6 | 76-80 | M | 1.3 | 1.7 | 6.3 | BUC (WD) |  | P | N | N | N |
| 7 | 46-50 | F | 0.6 | 3.0 | 9.9 | BUC, MTX |  | P | N | N | N |
| 8 | 61-65 | F | 0.8 | 3.0 | n.a. | BUC, NSAIDs |  | P | N | N | N |
| 9 | 76-80 | F | 1.3 | 2.5 | 13.0 | BUC (WD), PSL |  | P | N | N | N |
| 10 | 61-65 | M | 0.8 | 1.5 | n.a. | BUC (WD), PSL, MZ |  | P | N | N | N |
| 11 | 56-60 | F | 0.5 | 1.7 | 5.9 | BUC (WD), PSL, NSAIDs |  | P | N | N | N |
| 12 | 66-70 | M | 1.2 | 1.6 | 14.6 | BUC, SASP, NSAIDs | HBV(+) | P | N | N | N |
| 13 | 36-40 | F | 0.5 | 2.6 | 4.8 | BUC->ACT, NSAIDs, MZ, MTX |  | P | N | N | N |
| 14<sup>a</sup> (MN12) | 71-75 | M | 1.3 | 3.3 | 4.4 | BUC->AZA, PSL |  | P | N | N | N |
| 15 | 71-75 | F | 0.6 | 3.4 | 0.9 | BUC->Etanercept |  | P | N | N | N |
| 16 | 66-70 | F | 1.1 | 2.7 | 1.7 | BUC->GST, PSL | HCV(+) | P | N | N | N |
| 17<sup>a</sup> (MN11) | 66-70 | M | 0.8 | 0.7 | 10.2 | BUC->IGU, PSL |  | P | N | N | N |
| 18 | 71-75 | F | 1.0 | 2.7 | n.a. | GST (WD), PSL |  | P | N | N | N |
| 19 | 51-55 | F | 0.5 | 3.6 | 4.7 | MTX, PSL, NSAIDs |  | P | N | N | N |
| 20 <sup>a</sup> (MN13) | 56-60 | F | 1.1 | 1.7 | 12.7 | MTX->IGU | Colon cancer | P | N | N | N |
| 21 | 51-55 | F | 0.6 | 2.1 | n.a. | n.a. |  | P | N | N | N |
| 22 | 76-80 | F | 0.8 | 1.8 | n.a. | n.a. |  | P | N | N | N |
| 23 | 61-65 | F | 0.5 | 1.1 | 2.7 | n.a. |  | P | N | N | N |
| 24 | 66-70 | M | 0.7 | 2.1 | 9.2 | n.a. |  | P | N | N | N |
| 25 | 71-75 | F | 0.7 | 1.8 | n.a. | NSAIDs |  | P | N | N | N |
| 26 | 71-75 | F | 0.8 | 2.3 | n.a. | PSL |  | P | N | N | N |
| 27 | 76-80 | F | 0.7 | 3.2 | n.a. | PSL, MTX |  | P | N | N | N |
| 28 | 61-65 | F | 0.6 | 2.1 | n.a. | ACT |  | N | P | N | N |
| 29 | 66-70 | F | 0.9 | 3.3 | 10.1 | MTX, PSL |  | N | P | N | N |
| 30 | 56-60 | F | 1.0 | 3.4 | n.a. | ACT, GST, NSAIDs |  | N | N | N | N |
| 31 | 71-75 | F | 0.6 | 2.6 | 9.8 | ACT, PSL | Breast cancer, MF | N | N | N | N |
| 32 | 66-70 | F | 4.2 | 2.5 | n.a. | BUC (WD) |  | N | N | N | N |
| 33 | 71-75 | F | 0.9 | 3.0 | 3.0 | BUC (WD), NSAIDs | MCTD, SS | N | N | N | N |
| 34 | 61-65 | M | 1.2 | 2.0 | 4.8 | PSL, CyA |  | N | N | N | N |

Abbreviations: ACT, actarit; AZA, azathioprine; BUC, bucillamine; Cr, creatinine; CyA, cyclosporine A; EXT1, exostosin 1; F, female; GST, gold thiomalic acid sodium; HBV, hepatitis B virus; HCV, hepatitis C virus; IGU, iguratimod; M, male; MCTD, mixed connective tissue disease; MF, myelofibrosis; MTX, methotrexate; MZ, mizoribine; N, negative; n.a., not available; NELL1, neural epidermal growth factor-like 1; NSAIDs, non-steroidal anti-inflammatory drugs; P, positive; PLA2R, M-type phospholipase A2 receptor; PSL, prednisolone; RA, rheumatoid arthritis; SASP, salazosulfapyridine; SS, Sjögren's syndrome; THSD7A, thrombospondin type-1 domain-containing 7A; WD, withdrawal
<sup>a</sup>Three cases from MN cohort
<sup>b</sup>Patient ages are listed with a range of 5 years to protect personal information.

The IHC results of the four MN antigens in the RA-MN cohort are shown in Figure 1, Table 3. Twenty-seven cases were NELL1 positive (79%) (cases 1-27), 2 cases were PLA2R positive (6%) (cases 28 and 29), and 5 cases were quadruple negative (15%) (cases 30-34). In the RA-MN cohort, the male-to-female ratio was 7:27, and the mean age was 64.9 years (SD±10.3). The mean serum creatinine level was 0.9 mg/dl (SD±0.6), the mean serum albumin level was 2.4 g/dl (SD±0.7), and the mean urinary protein was 7.2 g/gCr (SD±4.2). Table 3 shows the history of anti-rheumatic drug use as noted on the biopsy request form. The most commonly used drug was BUC in 19 cases (56%) (cases 1 to 17 and cases 32 and 33). This was followed by prednisolone in 13 cases (38%), non-steroidal anti-inflammatory drugs in 10 cases (29%), MTX in 6 cases (18%), actarit in 4 cases (12%), gold sodium thiomalate (GST) in 3 cases (9%), mizoribine in 2 cases (6%), iguratimod in 2 cases (6%), salazosulfapyridine in 1 case (3%), CyA in 1 case (3%), etanercept in 1 case (3%) and azathioprine in 1 case (3%). Four patients (3%) had an unknown drug use history. Because anti-rheumatic drugs were suspected as the cause of proteinuria, at the time of renal biopsy, 10 patients (29%) (9 BUC, 1 GST) were on withdrawal from the drugs and 6 patients (18%) (5 BUC, 1 MTX) had been switched to other medications.

Other complications included 2 cases of hepatitis C virus carrier (cases 1 and 16), 1 case of hepatitis B virus carrier (case 12), 1 case of colon cancer (case 20), 1 breast cancer complicated with myelofibrosis (case 31), and 1 case of mixed connective tissue disease complicated with Sjögren’s syndrome (case 33).

### Representative case of NELL1 and PLA2R-double-positive MN

The histopathological findings and results of serum antibody analysis for a case of NELL1 and PLA2R-double-positive MN (MN cohort case 16) are shown in Figure 2 and Supplementary Figure S1. Since his late 40s, he has been treated by his primary care physician for T2DM. In his early 50s, he had positive urinary protein and occult blood in urine. Then, he was referred to our nephrology department, where a renal biopsy was performed. In the renal biopsy specimen, light microscopy showed mild glomerular basement membrane thickening, immunofluorescence showed IgG deposits along the glomerular basement membrane in a granular pattern, and electron microscopy showed subepithelial and intramembranous electron dense deposits with spike formation, leading to a diagnosis of stage II MN (Figures 2A, B, E). He was then treated conservatively with antihypertensive and antiplatelet drugs. His urinary protein level stayed at 0.5∼1.0 g/gCr and his serum creatinine level stayed at sCr 0.9∼1.0 mg/dl. In his late 60s, he had increased urinary protein and decreased serum albumin. A recurrence of MN was suspected and a second biopsy was performed. In the second biopsy specimen, light microscopy showed thickening of the glomerular basement membrane, immunofluorescence showed IgG deposits in the glomerular basement membrane, and electron microscopy showed intramembranous electron lucent deposits and subepithelial electron dense deposits. Immunohistochemically, PLA2R was globally positive on glomeruli, and NELL1 was positive on glomeruli in a segmental pattern. Thus, a diagnosis of PLA2R, NELL1-double-positive MN (stage I-III) was made (Figures 2F-J). From the serum collected at the time of the second biopsy, the anti-NELL1 antibody was positive by Western blotting (Supplementary Figure S1), and anti-PLA2R antibody was positive by ELISA (index value: 1.1 (reference value: 1.0 or higher indicating positive). In addition, IHC for PLA2R and NELL1 was performed using FFPE tissue of the first renal biopsy performed in his early 50s, and the results were positive for PLA2R (Figure 2C) and negative for NELL1 (Figure 2D). These results suggest that NELL1-associated MN was added to the underlying PLA2R-associated MN, resulting in MN that is double positive for PLA2R and NELL1

**Figure 2.**
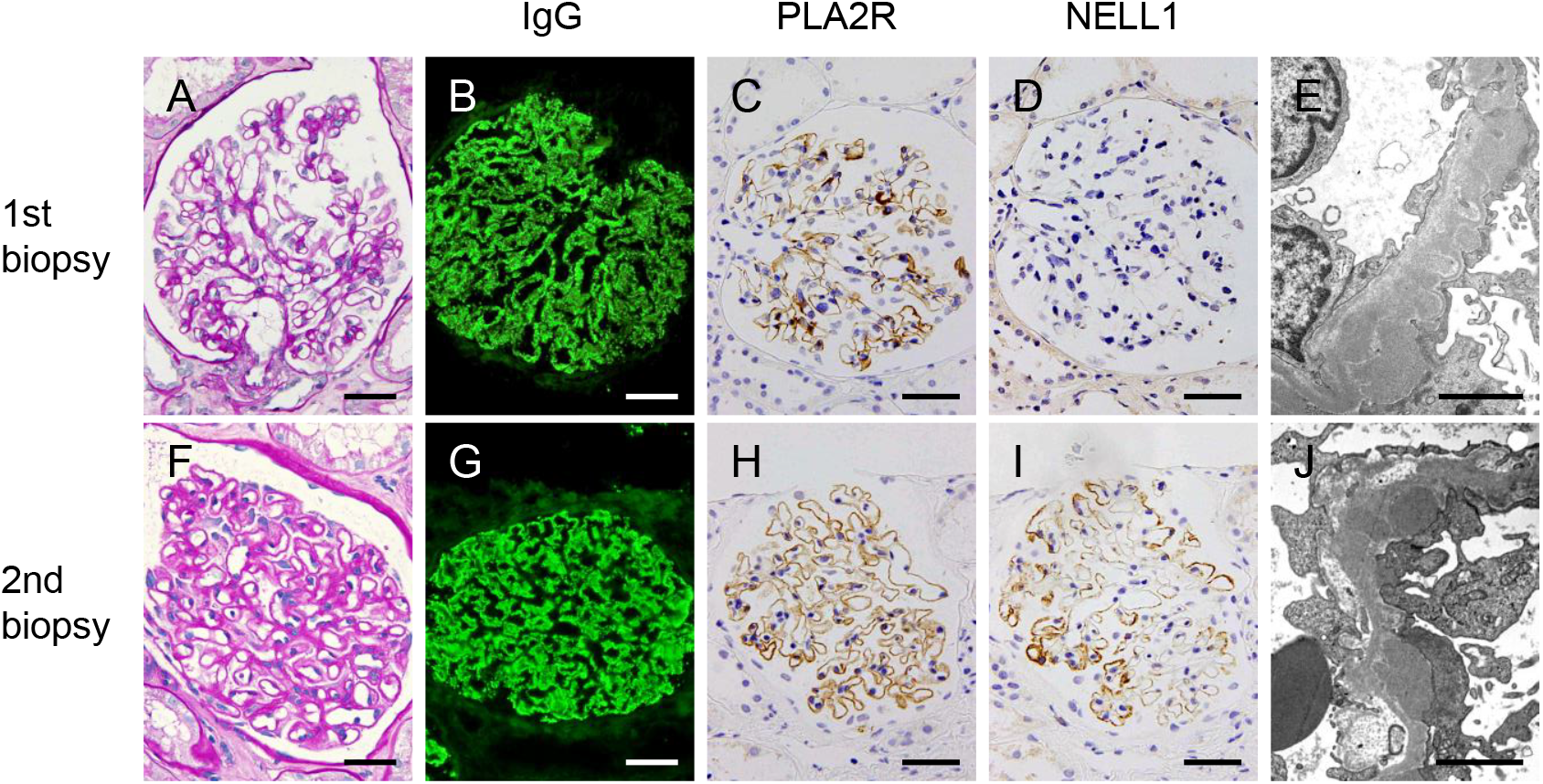
Histological picture of a case of PLA2R and NELL1-double-positive membranous nephropathy. Histological picture at the time of the first renal biopsy with diagnosis of membranous nephropathy (A-E) and second renal biopsy at the time of recurrence of membranous nephropathy (F-J) in case 16 of the MN cohort. (A, F) periodic acid-Schiff staining; (B, G) immunoglobulin G staining by immunofluorescence; (C, H) M-type phospholipase A2 receptor (PLA2R) staining by IHC; PLA2R was positive in the glomerulus in both the first and second biopsies. (D, I) neural epidermal growth factor-like 1 protein (NELL1) staining by IHC; NELL1 was negative in the first biopsy and positive in the second biopsy in a segmental pattern. (A-D, F-I) Bars = 50 μm (E, J) electron microscopy, Bars = 2 μm.

## Discussion

In the MN cohort, PLA2R was the most common antigen (40%) among the 4 antigens studied (PLA2R, NELL1, THSD7A, and EXT1). The frequency of PLA2R-associated MN among primary MN cases was reported to be higher in Europe, the United States, and China, accounting for 70-94% of MN cases ^1,14,15^. The frequency of PLA2R-associated MN among primary MN cases in Japan was reported to be about 53%, which is considerably less frequent than in Europe, US, or China ^16,17^. The frequency of PLA2R-associated MN in our MN cohort was slightly lower than the previously reported frequency of PLA2R-associated MN in primary MN cases in Japan. This may be due to the inclusion of secondary MN other than lupus nephritis type V in the MN cohort and the regional characteristics of the patients.

In the MN cohort, NELL1 was the second most common antigen after PLA2R and was found in 13% of patients with MN. It is widely recognized that NELL1-associated MN is often secondary MN. The most well-known association is with malignancy, and up to 33% of NELL1-associated MN cases are thought to be related to malignancy ^18^. Other known background conditions for NELL1-associated MN include associations with the intake of alpha lipoic acid^19^, intake of traditional indigenous medicine ^20^, hematopoietic stem cell transplantation ^21^, and post-kidney transplantation ^22^. In our MN cohort, 2 of the 16 patients (12.5%) with NELL1-associated MN had colon cancer, supporting the previously reported association with malignancy.

Patients with NELL1-associated MN in the MN cohort included 5 patients with rheumatic diseases (3 with RA, 1 with PMR, and 1 with SpA), 3 of whom (2 with RA and 1 with SpA) were treated with BUC. To our knowledge, no association between rheumatic diseases or BUC use and NELL1-associated MN has been reported, which led us to plan the next RA-MN cohort. It should be added that there was a recent report from Japan that 2 out of 4 patients with NELL1-associated MN were RA patients^17^.

The RA-MN cohort had a high rate of NELL1-associated MN (79%). In addition, 56% of the patients in the RA-MN cohort were using BUC. BUC is a thiol compound containing two thiol groups that was developed in Japan and has a structure similar to that of D-penicillamine. Like D-penicillamine, it is known to cause MN as an adverse effect. BUC-associated MN develops with proteinuria 2-11 months after the initiation of BUC therapy, and the proteinuria resolves with discontinuation of the drug in most cases ^23^. The histopathology most often shows stage I MN ^23,24^. There has been no report showing an association between BUC or D-penicillamine and NELL1-associated MN.

Although the mechanism of BUC-associated MN has remained unknown, reports of drug-induced pemphigus and insulin autoimmune syndrome (IAS), known as autoimmune diseases caused by thiol compounds, may be helpful in analogizing the mechanism of BUC-associated MN. In drug-induced pemphigus, thiol compounds such as D-penicillamine, captopril, and BUC are known to be causative agents ^25-27^. As the pathogenesis of drug-induced pemphigus caused by thiol compounds, it has been postulated that thiol compounds react with thiol groups that are abundant in the spinous and granular layers of the epidermis, causing acantholysis and subsequent production of autoantibodies against the pemphigus antigens ^27,28^.

IAS is a condition in which anti-insulin antibodies are produced in the absence of prior treatment with insulin, and the anti-insulin antibodies repeatedly bind to and release autologous insulin, resulting in hypoglycemia and postprandial hyperglycemia^29^. The reason that IAS is more common in Japan and less common in the West is thought to be that DRB1*406, an HLA-DR4 allele common in the Japanese population, is associated with IAS susceptibility^30^. In Japan, 41% of IAS cases occur in association with thiol compounds such as captopril, D-penicillamine, alpha lipoic acid, and BUC ^29,31^. A possible mechanism of IAS caused by thiol compounds is that thiol compounds interact with the disulfide bond sites of the insulin molecule, changing the three-dimensional structure of the insulin molecule, increasing its immunogenicity and inducing the production of autoantibodies^32^.

Thus, it is assumed that in thiol compound-induced autoimmune diseases, the thiol-disulfide exchange reaction between the thiol compounds and thiol groups or disulfide bond sites in the target protein results in structural changes in the target protein, leading to the production of autoantibodies.

Then, what mechanisms might be postulated in BUC-induced NELL1-associated MN? Human NELL1 is a protein with several structural motifs, including a thrombospondin module 1 domain-like domain, a coiled-coil domain, four von Willebrand factor type C domains, and six epidermal growth factor-like domains ^33^. Among these, the coiled-coil domain involved in oligomer formation, the von Willebrand factor type C domain involved in binding to bone morphogenetic protein, and the C-terminus involved in binding to integrin are rich in cysteine residues, suggesting the existence of multiple inter- and intra-molecular disulfide bond sites in the NELL1 protein ^34^. Thus, NELL1 is a potential target of thiol compounds and NELL1-associated MN induced by BUC may be caused by a mechanism that is similar to that of drug-induced pemphigus and IAS. Since alpha lipoic acid is also a thiol compound, it is possible that NELL1-associated MN due to alpha lipoic acid intake also occurs by the similar mechanism as presented here.

The RA-MN cohort also included NELL1-associated MN cases that were not treated with BUC (cases 18, 19, 20, 25, 26). These cases suggest that drugs other than BUC may be responsible for NELL1-associated MN or that RA itself may induce NELL1-associated MN. We await the accumulation of future cases to elucidate the mechanism of NELL1-associated MN in RA patients not treated with BUC.

In addition, 10 of 16 patients (63%) in the MN cohort had T2DM. This frequency of T2DM was much higher than the prevalence of T2DM (23.5% in men and 12.03% in women; 2020 data) in the Japanese population aged 70 years or older, the prevalent generation of NELL1-associated MN ^35^. In general, renal biopsies performed in T2DM patients often reveal MN as a non-diabetic glomerular disease, with 8.2% of patients in the United States and 21.8% in Japan reported to have MN ^36,37^. In addition, a Swedish epidemiological study revealed that patients with T2DM are at higher risk of developing various autoimmune diseases, and a chronic inflammation status, which exists in the background of both T2DM and autoimmune diseases, has been postulated as a common pathogenesis for both conditions ^38^. Although it will be difficult to clarify the causal relationship between T2DM and NELL1-associated MN, which is caused by multiple environmental and genetic factors, we hope that these relationships will be explained in future subsequent reports from other regions.

Since the discovery of PLA2R and the subsequent discovery of multiple MN-specific antigens, a small number of cases of MN are known to be simultaneously positive for both antigens, with PLA2R and THSD7A being the most frequently reported ^39-42^. Still, there are also reports of PLA2R and NELL1 double-positive cases ^14^. It is notable that in case 16 of the MN cohort, biopsies were performed twice and the first biopsy showed MN with a single positive for PLA2R, but the second biopsy showed MN with a double-positive for PLA2R and NELL1. In this case, the presence of anti-PLA2R and anti-NELL1 antibodies was found in serum at the time of the second biopsy, suggesting the possibility that a single antibody recognizes both PLA2R and NELL1 epitopes or that anti-PLA2R and anti-NELL1 antibodies are present separately. Since no homology between PLA2R and NELL1 amino acid sequences was confirmed by a BLAST search, it is unlikely that a single antibody cross-reacts with PLA2R and NELL1. Therefore, in case 16 of the MN cohort, we considered it highly likely that anti-PLA2R and anti-NELL1 antibodies are present separately. In chronic autoimmune diseases, it is often observed that a variety of autoantibodies are produced during the course of the disease, and epitope spreading is one of the mechanisms by which this occurs. Epitope spreading is a phenomenon in which, in the antibody production response, specific antibodies are initially produced to a particular epitope, but gradually antibodies are produced to epitopes other than the initial epitope, resulting in diversity in antibody-antigen recognition. Two types of epitope spreading are known: intramolecular epitope spreading, in which antibodies are produced against another epitope in the same molecule, and intermolecular epitope spreading, in which antibodies are directed against an epitope in another molecule ^43^.

Intramolecular epitope spreading has been reported in PLA2R-associated MN. According to that report, some patients who initially had antibodies recognizing the cysteine-rich domain of PLA2R had antibodies recognizing C-type lectin domain 1 or C-type lectin domain 7 at the time of relapse ^44^. Although intermolecular epitope spreading in PLA2R-associated MN has not yet been reported, we speculate that the intermolecular epitope spreading to the NELL1 molecule in PLA2R-associated MN may have occurred in this case. In the future, the epitope transition of antibodies can be examined in similar cases from before the onset of the disease, a better understanding of the pathogenesis of double antigen-positive MN should be achieved.

The limitations of this study are as follows. The study was conducted at a single center in Japan, was limited to Asians, and the patient population was almost restricted to the Hokkaido area. It is unclear whether the results can be generalized to other ethnic groups or regions. In addition, the RA-MN cohort included many historical cases for which there were no medical records, and clinical information in the RA-MN cohort was limited to the content of the kidney biopsy order form, which may not be sufficient.

In conclusion, in the MN cohort, 4 antigens (PLA2R, NELL1, THSD7A, and EXT1) were examined in patients with MN in Japan, and NELL1 was found to be the second most common antigen after PLA2R with a frequency of 13%. NELL1-associated MN was associated with malignancy (12.5%) as well as rheumatic disease with BUC use (18.8%) and T2DM (63%). In the RA-MN cohort, among MN patients with RA, 79% of the patients had NELL1-associated MN and 56% of the patients were using BUC.

The results indicate that NELL1-associated MN is associated not only with malignancies but also with BUC use for RA and T2DM.

## Data Availability

All data produced in the present study are available upon reasonable request to the authors

## Acknowledgements

We are grateful to the Pathology Section, Department of Clinical Laboratory, Sapporo City General Hospital for routinely preparing excellent renal biopsy specimens. We are grateful to Mr. Masaki Kutsumura, medical secretary, for his great help in data collection. We thank Dr. Shin’ichi Akiyama, Division of Nephrology, Department of Internal Medicine, Nagoya University Graduate School of Medicine, for useful discussions on epitope spreading in membranous nephropathy.

