## Supplementary Figure S1 for "Bucillamine use for rheumatoid arthritis and type 2 diabetes mellitus are associated with neural epidermal growth factor-like 1 (NELL1)-associated membranous nephropathy"

Supplementary Figure S1. Detection of anti-NELL1 antibodies in serum using recombinant human NELL1 protein. Under a non-reducing condition, a 420-kDa band corresponding to the homotrimer of NELL1 protein was demonstrated by Western blotting in serum at the time of the second renal biopsy of case 16 of MN cohort but not in serum from patients with IgA nephropathy (IgAN) and minimal change disease (MCD).

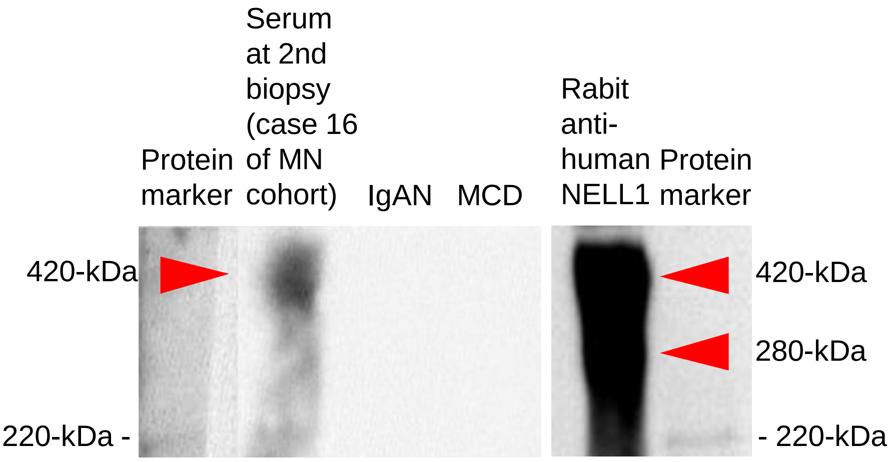
