## Supplementary Methods for "Bucillamine use for rheumatoid arthritis and type 2 diabetes mellitus are associated with neural epidermal growth factor-like 1 (NELL1)-associated membranous nephropathy"

**Histological analysis**

Specimens for light microscopy, immunofluorescence, and electron microscopy were prepared by methods commonly used. For light microscopic analysis, FFPE tissue was thinly sliced to 3 µm in thickness, and hematoxylin and eosin, periodic acid-Schiff, elastica-Masson trichrome, and periodic acid methenamine silver staining were performed. In the MN cohort, total and sclerosing glomeruli were counted, the degree of interstitial fibrosis and tubular atrophy (IFTA) in the cortical area was assessed in 5% increments, and the presence of mesangial proliferative changes in the glomeruli was evaluated.

**Immunohistochemical analysis**

We studied four specific antigens for MN (PLA2R, THSD7A, NELL1, and EXT1) by IHC. Three-µm-thick FFPE sections were prepared and stained using an automated immunostaining system (Leica BOND III and BOND MAX) based on the enzyme-labeled polymer method. For detection of PLA2R, NELL1, and EXT1, the BOND Enzyme Pretreatment Kit (Leica) was used at a dilution of 1:20 for antigen retrieval at 37°C for 5 minutes. Primary antibodies used and their concentrations were anti-PLA2R1 antibody (rabbit polyclonal, HPA012657, Sigma-Aldrich, 1:200), anti-NELL1 antibody (rabbit polyclonal, ab197315, Abcam, 1:100) and anti-EXT1 antibody (rabbit polyclonal, ab126305, Abcam, 1:100). For detection of THSD7A, antigen retrieval was performed using BOND Epitope Retrieval Solution 2 (pH 9.0, Leica) at 100°C for 20 minutes. The primary antibody used and its concentration was anti-THSD7A antibody (rabbit polyclonal, HPA000923, Sigma-Aldrich, 1:800). Primary antibody reaction conditions for all antigens were 15 min at room temperature, and detection was performed with BOND Polymer Refine Detection (Leica).

**Immunofluorescence analysis**

Frozen sections of 3 µm in thickness were prepared and subjected to direct immunofluorescence by conventional methods. In the MN cohort, IgA, IgM, IgG, C3, C1q, κ, λ, IgG1, IgG2, IgG3, and IgG4 were subjected to direct immunofluorescence using fluorescein isothiocyanate-labeled antibodies. Primary antibodies used and their concentrations were anti-human IgA antibody (rabbit polyclonal, Dako, F0316, 1:20), anti-human IgM antibody (goat polyclonal, MP Biomedicals, 55153, 1:80), anti-human IgG antibody (rabbit polyclonal, Dako, F0315, 1:20), anti-human C3 antibody (goat polyclonal, MP Biomedicals, 55167, 1:40), anti-human C1q antibody (goat polyclonal, MP Biomedicals, 55166, 1:20), anti-human κ-light chain antibody (rabbit polyclonal, Dako, F0198, 1:40), anti-human λ-light chain antibody (rabbit polyclonal, Dako, F0199, 1:80), anti-human IgG1 antibody (sheep polyclonal, Binding Site, AF006, 1:10), anti-human IgG2 antibody (sheep polyclonal, Binding Site, AF007, 1:20), anti-human IgG3 antibody (sheep polyclonal, Binding Site, AF008, 1:20) and anti-human IgG4 antibody (sheep polyclonal, Binding Site, AF009, 1:10). The reaction conditions for the primary antibodies were 37°C for 60 minutes. Washing was performed twice with phosphate-buffered saline (PBS) for 5 minutes, and after sealing with glycerol, the slides were subjected to observation under a fluorescence microscope. Fluorescence intensity was evaluated on a four-level scale from 0 to 3+ (0, negative; 1+, weakly positive; 2+, moderately positive; 3+, strongly positive).

**Electron microscopic analysis**

In the MN cohort, staging of MN was performed with Ehrenreich and Churg classification in 10 cases that were analyzed by electron microscopy.

**Detection of anti-NELL1 and anti-PLA2R antibodies in patient serum**

Detection of anti-NELL1 antibodies in patient sera was performed by Western blotting according to the previously described method^4^. Sodium dodecyl sulfate (SDS)-polyacrylamide gel electrophoresis under non-reducing conditions was performed on a 6% SDS-polyacrylamide gel with 0.5 µg of NELL1 recombinant protein (R&D Systems) per well applied and transferred to a polyvinylidene difluoride membrane. Blocking was performed in PBS-tween buffer containing 1% skim milk at room temperature for 1 hour. Patient serum was diluted at 1:50 and rabbit anti-human NELL1 antibody (Abcam) was diluted at 1:500 for the primary antibody reaction. Primary antibody reactions were performed at 4°C overnight. Secondary antibody reactions were performed at room temperature for 2 hours. Horseradish peroxidase (HRP)-conjugated rabbit anti-human IgG antibody (GeneTex: 1:10000) and HRP-conjugated goat anti-rabbit IgG antibody (Jackson ImmunoResearch: 1:20000) were used as secondary antibodies. Immunostar LD (Fujifilm Wako Pure Chemicals) was used for chemiluminescence.

Detection of anti-PLA2R antibodies in patient sera was performed by a commercial laboratory (BML, INC.) that provides anti-PLA2R antibody testing using an ELISA kit manufactured by EUROIMMUN.
